# Outbreak of SARS-CoV-2 B.1.617.2 (Delta) variant in a Nursing Home 28 weeks after two doses of mRNA anti-Covid-19 vaccines: evidence of a waning immunity

**DOI:** 10.1101/2021.10.25.21265370

**Authors:** Alice Pierobon, Alessandra Dal Zotto, Antonio Antico, De Antoni Mario Ernesto, Liviano Vianello, Monica Gennari, Antonio Di Caprio, Francesca Russo, Gianfranco Brambilla, Mario Saugo

## Abstract

**Objectives:** Description of a SARS-CoV-2 B.1.617.2 (Delta) variant outbreak among residents (N=69) and Health Workers (HWs: N=69) of a small Nursing Home in Northern-East Italy, with full vaccination coverage of 91 and 82%, respectively. Evaluation of the Anti-Spike IgG titers 28 weeks after the mRNA vaccine boosts against SARS-COV-2 infection and severe Covid-19.

**Materials and methods:** A timely collection of sera within 48h from the index case; anti-Spike IgG determination (expressed as Binding Antibody Units – BAU/mL) through a commercial quantitative assay; SARS-CoV-2 diagnostics via RT-PCR, and full-genome sequencing for lineage characterization. Residents were grouped according to anti-Spike IgG titers (≤ 50, 51-1000, and > 1000 BAU/mL) and resulting protection against the infection and the severe disease was measured.

**Results:** 0/20 HWs and 14/59 (24%) residents fully vaccinated and without a previous SARS-CoV-2 infection showed anti-Spike IgG lower/equal to 50 BAU/mL (1-sided Fisher exact p=0.011). Among these residents, a level of anti-Spike IgG ≤50 BAU/mL resulted in a higher risk of SARS-CoV-2 infection (RR=1.55, CI95% 1.17-2.05) and severe Covid-19 disease (RR=5.33, CI95% 1.83-15.57). Conclusion Low levels of SARS-CoV-2 neutralizing anti-Spike IgG in serum 28 weeks after the administration of the second dose parallels the waning of vaccine protection.

## Introduction

Outbreaks in Long-Term Care Facilities (LTCF) are considered a sentinel event for re-infection after full vaccination, due to a sub-optimal antibody response due to the age of residents, underlying comorbidities, and ongoing corticosteroids therapy [1]. In this regard, the recent spread of B.1.617.2 SARS-CoV-2 Variant of Concern (VOC Delta) is of particular concern [2, 3, 4].

In this paper, we report an outbreak in a small nursing home in Northern-East Italy, started on the end of August 2021, along with an extensive and timely serological dosage of the SARS-CoV-2 anti-Spike IgG among residents. The aim is to shed some light on how anti-Spike IgG titer decrease parallels the waning of m-RNA vaccine protection against the VOC Delta.

## Materials and Methods

On the end of August 2021, a Healthcare Worker (HW) tested positive to routine checks performed via a microfluidic immunofluorescence assay for the qualitative detection of nucleocapsid antigens (LumiraDX SARS-CoV-2 Ag).

Immediately, all residents and the nursery staff were screened with the same assay, and 12 additional residents and 5 HWs tested positive. Within the next 48h, confirmatory molecular RT-PCR tests on nasopharingeal swabs were carried out among all residents and HW staff and 10 swabs were sent to the accredited laboratory of Istituto Zooprofilattico Sperimentale delle Venezie for the Sars-CoV-2 B.1.617.2 lineage characterization via Illumina MiSec platform. At the same time, blood samples were drawn from residents and a sample of HWs, for the determination of both the anti-Spike IgM and IgG in sera (Abbott SARS-CoV-2 IgG II Quant Assay). IgG results were expressed in Binding Antibody Units (BAU) per mL, using the manufacturer’s conversion factors, based on the WHO International Standard Anti-SARS-CoV-2 Immunoglobulin (NIBSC code 20-136) [5]. Testing was carried out at the Local Health Unit (ULSS 7 Pedemontana) analytical lab, under QA/QC. The following two conventional threshold levels of anti-Spike IgG were adopted: ≤50 BAU/mL and >1000 BAU/mL.

The outbreak lasted 42 days, accounting for 10 day after the last SARS-CoV-2 infection. Affected residents were grouped as follows: A) Asymptomatic: including < 38 °C fever or headache; B) Symptomatic: > 38 °C fever, cough, diarrhea, anorexia, lethargy, psychomotor retardation, mental confusion; and, C) Severe: dyspnoea, desaturation < p92 leading to fatal outcomes. Previous immune stimuli were considered: doses of mRNA Covid-19 vaccination received within the first decade of August and SARS-CoV-2 RT-PCR-confirmed infection before the end of May, 2021.

The relationship between anti-Spike IgG titers and Covid-19 status among fully vaccinated residents without a previous SARS-CoV-2 infection was described through a tabular and graphical bivariate analysis. Cumulative Risk Ratios (RR) and vaccine effectiveness (VE) against infection and severe Covid-19 were calculated according to the formula: 1 - RR, where RR represents the cumulative risks of an adverse outcome comparing those with anti-Spike IgG ≤ 50 vs. > 50 WHO BAU/mL.

The study was approved by the competent USLL 7 Ethics Committee on the end of August, 2021.

## Results

On the index-case incidence date, among the 69 residents (80% female, 84% ≥80y aged, 88% completely dependent), and the 69 HWs present, 3 and 5 individuals, respectively, had a previous SARS-CoV-2 RT-PCR-confirmed infection (March 2020 - April 2021). The full vaccination coverage was 83% among HWs and 91% among residents, mainly via BNT162B2 m-RNA vaccine (98%), with the boost dose received on average 196±36 days before the outbreak. Among residents, 2 subjects received only the priming, and 4 resulted unvaccinated; among HWs, 6 and 6.

Within 30 days, 46 and 14 new cases were found among residents and HWs, respectively (cumulative incidence: 67%; 20%), with 75% of cases within the first week from the Index case. Spike sequencing on 7 residents and 3 HWs swabs always confirmed the VOC Delta lineage.

Among residents, 13 individuals were asymptomatic, 21 symptomatic, and 12 severe Covid-19 cases (8 deaths). Two deaths (*ab ingestis* pneumonia and cachexia), occurring 11 days from the last negative PCR-test, were not accounted as fatal Covid-19 cases.

Anti-Spike IgG geometric mean was 161 (CI95% 98-268) and 625 (CI95% 289-1353) BAU/mL among tested residents (66/69) and HWs (23/66), respectively; residents ≤80y vs. >80y showed a geometric mean of 585 (CI95% 263-1300) and 103 (CI95% 58-186) BAU/mL, respectively.

Among tested fully vaccinated and without previous SARS-CoV-2 infection individuals, 0/20 HWs, but 14/59 (24%) tested residents had anti-Spike IgG ≤50 WHO BAU/mL (1-sided Fisher exact p= 0.011).

As for these residents, the elapsed time since the boost at the onset of the outbreak was 199 (interdecile range: 154-209) days, and an anti-Spike IgG titer ≤50 BAU/mL resulted in a higher risk of SARS-CoV-2 infection (RR=1.55, CI95% 1.17-2.05) and severe Covid-19 disease (RR=5.33, CI95% 1.83-15.57) (**Table 1, Figure 1**). In terms of VE, the corresponding figures against SARS-CoV-2 infection and severe Covid-19 were 35% (CI95% 15-51%) and 79% (CI95% 37-93%), respectively; close results were obtained including the unvaccinated (n=2) and partially vaccinated (n=3) residents into the comparison group (data not reported).

**Table 1.** SARS-CoV-2 infection and COVID-19 disease incidence, according to the threshold Anti-Spike IgG level (<= vs >50 BAU/mL) in 59 fully mRNA vaccinated residents.

| IgG anti-S | SARS-CoV-2 | Covid-19 |  |  | Total |
| --- | --- | --- | --- | --- | --- |
|  |  | Asymptomatic | Symptomatic | Severe |  |
| **$> 50$ BAU/mL** | **negative** |  |  |  |  |
| No | 1 (7%) | 3 (21%) | 4 (29%) | 6 (43%) | 14 (100%) |
| Yes | 18 (40%) | 9 (20%) | 14 (31%) | 4 (9%) | 45 (100%) |
| Total | 19 (32%) | 12 (20%) | 18 (31%) | 10 (17%) | 59 (100%) |

Fisher exact $p=0.012$

**Figure 1.**
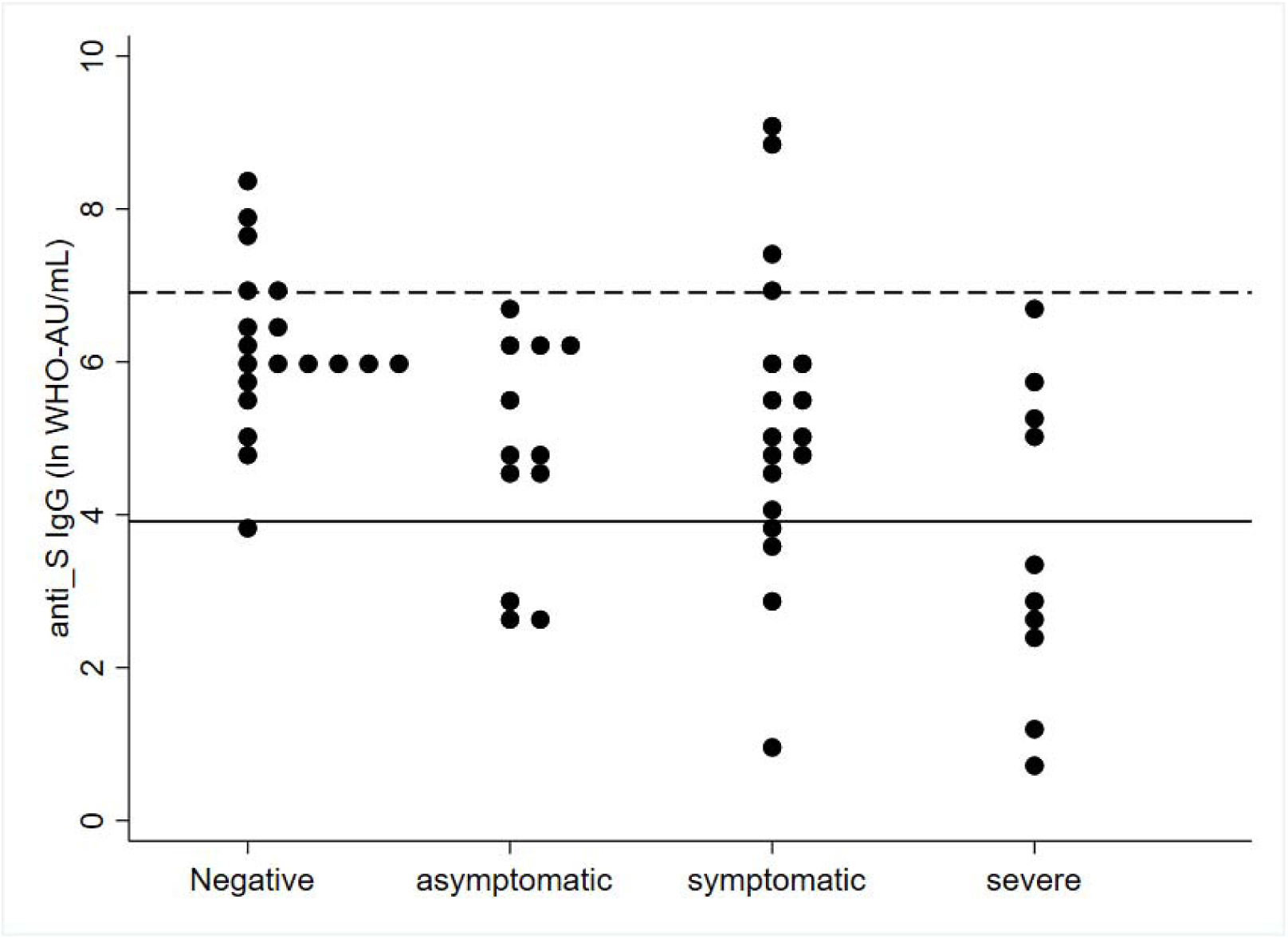
Cumulative Incidence of SARS-CoV-2 infection and Covid-19 disease among 59 residents fully vaccinated with mRNA vaccine 28 weeks before, and with no previous SARS-CoV-2 PCR-confirmed infection, by anti-S IgG level (BAU/mL) measured at the beginning of the outbreak. Full and dotted black lines represent two conventional threshold levels of 50 and 1000 BAU/mL

## Discussion

One-fourth of 59 fully vaccinated Nursing Home residents with no previous SARS-COV-2 infection residents had anti-Spike IgG ≤ 50 WHO BAU/mL about 28 weeks after the boost. This translates in a VE of 35% (CI95% 15-51%) against infection and 79% (CI95% 37-93%) against severe Covid-19 disease, suggesting that the protection conferred from an anti-Spike IgG titer ≤50 BAU/mL is very low, if any.

A large US-CDC study carried out among 3,862 nursing home residents fully vaccinated with m-RNA vaccines, reports a VE against SARS-CoV-2 infection of 53% (CI 95% 49-57%) comparing post-Delta (June 2-August 1, 2021) with pre-Delta period [6]. Otherwise, a nationwide UK study computed a VE of 71% (CI95% 41-86%) against Delta hospitalization among clinically extremely vulnerable ≥65y aged 20+ weeks after the BNT162B2 boost [7].

In its Rapid Risk Assessment on Covid-19 outbreaks in LTCFs, the European Center for Disease Prevention and Control (ECDC) reported 2/18 VOC Delta outbreaks until July 2021, with 63 fully vaccinated and 12 partially vaccinated or unvaccinated residents. The reported attack rate among fully vaccinated residents is 31/63 (49%), with 5/63 (8%) Covid-19 hospitalizations [4]. These figures are somewhat lower in comparison with our study, which also shows a higher vaccination coverage among residents and HWs; this is probably due to differences in elapsed time from the boost.

To this purpose, a recent study evaluated anti-S Ig Titer among 472 LTCF residents with Elecsys Anti-SARS-CoV-2 S Assay (April-May 2021), 64 days (IQR 63-65) from the BNT162b2 boost. The high, medium, low, null titer groups (>1,000, 101-1,000, 1-100 and <1 BAU/mL) showed frequencies of 62%, 22%, 16%, and 4%, respectively. The risk to be a non-responder was 8 times higher in individuals with no previous SARS-Cov-2 infection (about 30% of the tested residents) [1]. In the present case study, only 5 (7%) residents had a previous SARS-CoV-2 infection in the pre-Delta period; none of them got infected during this outbreak.

To conclude, this study firstly reports a VOC Delta outbreak in an Italian Nursing Home [8, 9]; the extensive and timely measurement of anti-Spike IgG among residents provides valuable immunological information. Due to the small size and the limits of our observation, further research is needed to define appropriate threshold anti-Spike IgG levels to identify and protect at best those residents at higher risk of severe Covid-19 from VOC Delta lineages.

Authors declare no competing interest

## Data Availability

All data produced in the present work are contained in the manuscript.
https://www.gisaid.org/; ID numbers: EPI_ISL_4556244; EPI_ISL_4556245; EPI_ISL_4556247; EPI_ISL_4556248; EPI_ISL_4556250; EPI_ISL_4556252; EPI_ISL_4556253; EPI_ISL_4556256; EPI_ISL_4556257, EPI_ISL_4556258.

## Acknowledgments

Authors are in debt to dr.ssa Isabella Monne of Istituto Zooprofilattico Sperimentale delle Venezie for the VOC Delta full genotyping available through the open access GIS AID repository (https://www.gisaid.org/), with the following ID numbers: EPI_ISL_4556244; EPI_ISL_4556245; EPI_ISL_4556247; EPI_ISL_4556248; EPI_ISL_4556250; EPI_ISL_4556252; EPI_ISL_4556253; EPI_ISL_4556256; EPI_ISL_4556257, EPI_ISL_4556258.

## Authors’ contributions

Conceptualisation: AA, FR, LV, ADC, MS; Investigation: AA, AP, MEDA, MG, ADZ; formal analysis: AP, ADZ, MS; writing: GB; supervision: MS.

